# The medical signature of musicians: A Phenome-wide association study using an Electronic Health Record database

**DOI:** 10.1101/2020.08.14.20175109

**Authors:** Maria Niarchou, George Lin, Miriam D. Lense, Reyna Gordon, Lea Davis

## Abstract

**Background:** A limited set of previous studies suggest musicians may be at higher risk for a unique set of medical and mental health problems. To address the limitations of previous studies, we examined trends in the medical care of musicians in Vanderbilt’s Electronic Health Record research database.

**Methods:** We mined clinical notes for a principled collection of keywords and regular expressions which indicated that the patient was a musician. We identified 9,803 “musicians” that we matched for sex, median age (across the medical record), ethnicity, race, length of record and number of visits with 49,015 controls. We fitted 1,263 logistic regression models (one for each phenotype classification).

**Results:** 257 medical diagnoses were significantly more prevalent in musicians than controls after strict Bonferroni adjustment for the total number of phenotypes tested (p-value<7.6 x 10^−6^). Diseases of the larynx and the vocal cords (OR=2.32, p<2.84 x 10^−191^), and hearing loss (OR=1.36, p=5.98 x 10^−97^) were among the top associations. Anxiety disorder (OR=1.25, p=7.67 x 10^−71^), and Major depressive disorder (OR=1.21, p=4.88 x 10^−36^) were also strongly associated with musician status. Fifteen phenotypes were significantly more prevalent in non-musicians than musicians, including Coronary atherosclerosis (OR=0.91, p=1.77 x 10^−10^), and Ischemic Heart Disease (OR=0.92, p=1.65 x 10^−09^).

**Conclusions:** Although being a musician was related to a large number of occupational health problems, we also identified protective effects of musicianship in which certain diagnoses were less common in musicians than in non-musicians, indicating that active musical engagement could have similar health benefits to athletic engagement.

**What is already known on this subject?:** Being a musician is a physically and psychologically demanding undertaking. Previous studies suggest musicians may be at higher risk for a variety of medical and mental health problems.

**What this study adds?:** Most studies of musicians’ health, are based on relatively small sample sizes, self-report questionnaire data, and often lack controls groups. In a sample of 14,379 musician cases and 71,895 matched controls identified in an Electronic Health Record database (EHR), we conducted the first and largest study to date to identify the medical diagnoses associated with musician patients in an EHR context. We replicated previous associations of musician status with medical problems and we identified a number of protective effects by observing diagnoses such as cardiovascular disease, respiratory failure, and renal failure, that were less common in musicians than in controls, in line with literature indicating that active musical engagement has similar health benefits to athletic engagement.

## Introduction

Similar to elite athletes, musicians undergo years of effortful training and dedicated practice in order to be able to play music at a professional level. The demands associated with the profession are enormous and can impact multiple aspects of a musician’s life and health. Lack of financial security and career stability, repeated competitive evaluations, and performance-related demands^1^ can be sources of stress while the physical demands to be able to perform at a high-level increases vulnerability to physical injuries (e.g., repetitive use injuries)^2^. Despite the challenges associated with this career path, the national infrastructure for professional musicians’ health is less robust than services for elite athletes, where sports-medicine strategies for improving wellness are the norm^3^, though there are notable exceptions of organizations like MusiCares (https://www.grammy.com/musicares/about) and a relatively small number of highly specialized medical clinics that specialize in musicians’ health.

Studies indicate a variety of physical health problems associated with being a musician, including musculoskeletal disorders^4^ (e.g., tendonitis)^5^, skin disorders^6^, respiratory problems^7^ and hearing loss^2^. Mental health problems are also common including performance-related anxiety^8^, anxiety, depression^9^, and sleep problems^10^. Alcohol, medication, and drug use are frequently encountered among recording musicians^11^ despite their known risks and contributions to the premature mortality of famous musicians (e.g., ^12^). At the same time, a study of 62 professional musicians indicated that the level of heart rate response during a concert performance was similar to the heart rate response of professional athletes, highlighting the high physiological demands of professional musicianship^13^. Most studies of musicians’ health, however, are based on relatively small sample sizes, self-report questionnaire data, and often lack control groups. Therefore, our understanding of the scope and prevalence of the physical and mental health problems of musicians is limited.

To address these limitations, we conducted an extensive study of 86,274 individuals to identify the health conditions for which musicians are more frequently coded, using data from Electronic Health Records (EHRs) in Vanderbilt University Medical Center’s de-identified research database. Nashville, known as “Music City, USA,” has a large population of working musicians, and Vanderbilt University Medical Center has a reputation for providing specialized medical care to musicians from the Nashville area and beyond. Our study examined the following questions: What are the most frequent physical and mental health problems of musicians reported in the EHR?, and: Given that there are sex differences in prevalence of many health problems, are there sex differences observed for any musician-associated health conditions? Given recent advances in phenome-wide association studies (PheWAS) methodologies for epidemiological discovery in data-rich electronic health record research databases^14^, we employed PheWAS approaches to conduct data-driven exploration in the sample. Taking into account that the health issues that musicians face can be related to their instrument^2^, we also examined what medical problems are associated with musicians who play different families of instruments (e.g., string instruments, brass instruments, voice). Finally, we also examined the health problems in children and adolescents, to explore potential musician-related health problems observed early in life.

## Materials and methods

### Vanderbilt’s Electronic Health Records (EHR) database

The study was conducted using the Synthetic Derivative (SD) database at Vanderbilt University Medical Center (VUMC). The SD is the de-identified version of the entire VUMC Electronic Medical Records system (EMR), which contains records of over 3 million unique individuals^15^. The database incorporates data from multiple components, including diagnostic and procedure codes, demographics, progress and nursing notes, problem lists, and medication histories. This study is approved by Vanderbilt’s Institutional Review Board for non-human subjects research (IRB #160302).

### The musician-related keyword selection

We determined the musician status in patient records through keyword searches in patients’ de-identified EHRs from the SD database. To generate an initial keyword list, we obtained the number of individuals per instrument type or musician-related profession from the Nashville Musicians Association (http://www.nashvillemusicians.org), the local affiliate of the American Federation of Musicians of the United States and Canada. The initial list included 100 keywords (Supplementary Table 1). Out of these, we selected the instruments/music-related professions that were endorsed by more than 10 individuals in the keyword list, which reduced the list to 44 keywords. We then conducted 300 chart reviews (7 charts per keyword), to examine whether the keywords are within context (i.e., whether they characterise individuals who are musicians). As a result of the searches, we removed eight keywords as they mostly captured non-musician related traits (e.g., arranger, contractor, copyist, programmer, recorder). Using only the name of the musical instrument returned results for which the keyword was unrelated to music contexts (e.g., bass fishing, accordion drainage bag, mandolin slicer, banjo curette), thus we restricted the search to 449 regular expressions (e.g., “play the”) plus the name of the musical instrument as a keyword (for details see Figure 1, Table 1, Supplementary Table 2), and four keywords that were added to the search separately (i.e., songwriter, musician, singer, vocalist). We also excluded from the search the following phrases as this was the name of a book for children with autism: “The Different Drummer”, “the different drummer”, “Different Drummer”, “different drummer”. We restricted our search to patients for whom VUMC was the most likely source of primary care using a heuristic “medical home” definition (i.e., five codes on different days over three years). This approach reduces false positive associations that could be due to systematic missingness in the data. The final search resulted in 14,929 individuals qualifying as musicians.

**Table 1.**
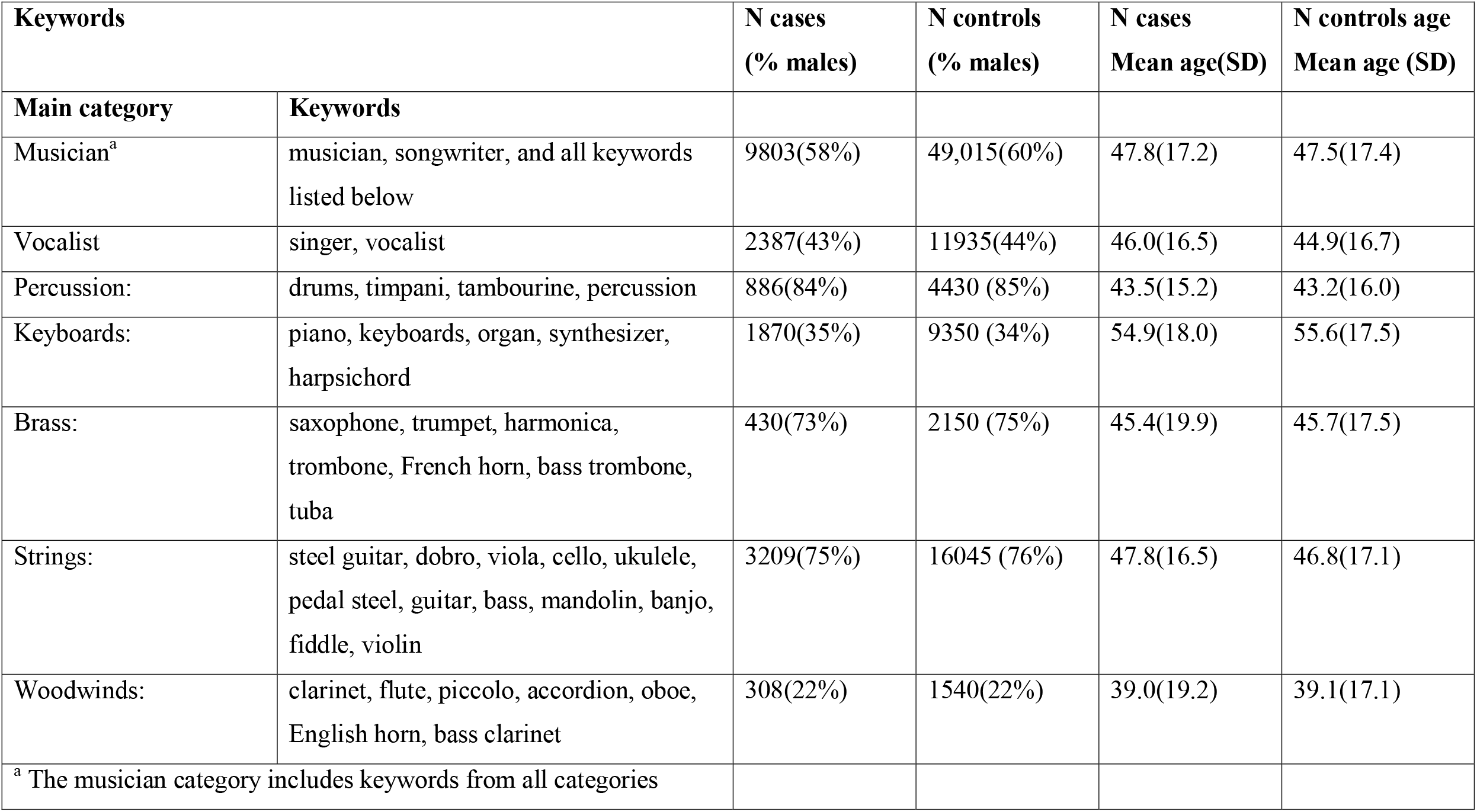
Keywords for instrument families and descriptives

**Figure 1.**
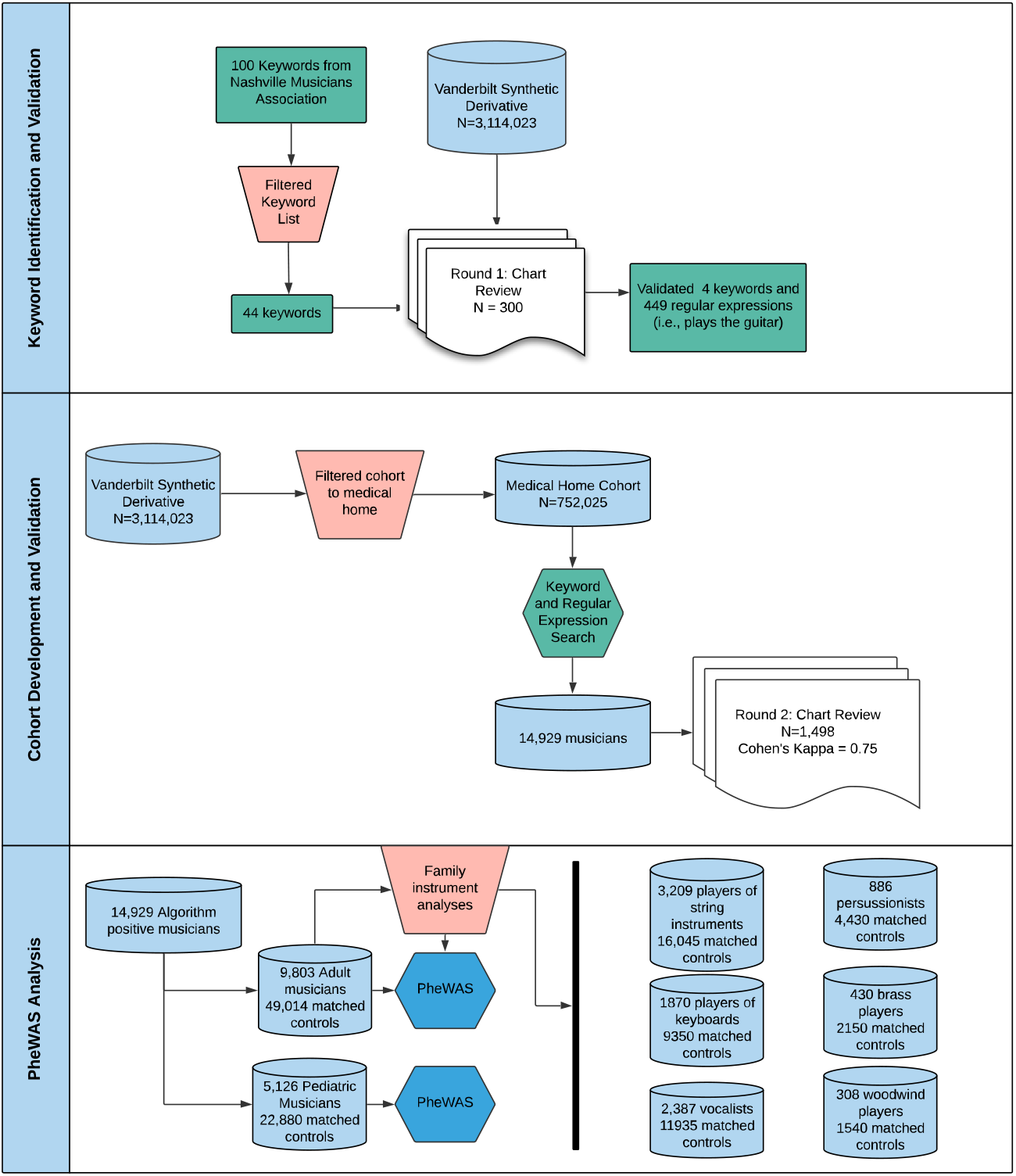
Keyword selection process and chart review

### Chart reviews

Two reviewers (authors MN and GL) each reviewed 10% (N=1,498) of randomly selected records from the set of musician cases identified by the keyword search described above, in order to establish the positive predictive value (PPV). Charts were evaluated for evidence of musicianship and did not restrict the search to professional musicians. Individuals were considered as musician cases if their record included evidence that they played a musical instrument or were a musician. The procedure was as following: each reviewer loaded the record of the individual in the SD interface and searched for the keyword in all the documents of the individual’s record. Each author separately read the phrase/paragraph within which this keyword was included and determined whether it was within the context of patient musician activity or not. The reviewer agreement was 97% based on 1,498 charts, and Kappa inter-rater reliability was 0.75 (substantial agreement). The approach yielded a high PPV of 93%.

For the control sample (i.e., non-musicians), we included all individuals that had no mention of musician keywords in their medical charts, and matched them to the musicians in a ration of 1:5 for median age across the medical record, length of record, number of visits, sex, race, and ethnicity. We used the R MatchIt package (using the nearest neighbour matching setting). Control (or non-musician) charts had no keywords from our initial list. It is possible that some controls include musicians who do not have record of these keywords in their chart. This type of misclassification (i.e., cases as controls) could result in reduced power, but would not result in increased type 1 error. Because of our large sample size, loss of power due to potential misclassification of cases as controls was not a significant cause of concern for our study.

### Statistical analyses

For our primary analyses, we restricted our sample to individuals with a median age of record 18 years of age and older. We performed a phenome-wide association study (PheWAS) ^16^, using EHR-derived case-control status for 1,263 phecodes (i.e., groups of medical diagnoses) using International Classification of Disease versions 9 and 10 (ICD-9 and ICD-10) codes and estimated associations with musician status (case versus controls), in Vanderbilt’s EHR database. ICD codes were converted to phecodes using the R PheWAS package ^14^. The mappings of the ICD codes to phecodes can be found here (http://phewascatalog.org). We restricted our analyses to phecodes that had over 100 cases or controls. We also repeated the approach to perform exploratory analyses for keywords within the family instrument categories (Table 1 & Supplementary Tables 4 to 9).

We conducted an additional set of analyses where we restricted our sample to children/adolescents (individuals with a median age of record between the ages of five and seventeen years old; Supplementary Table 10).

## Results

### Associations with musician status

For our primary analyses, we identified 9,803 adult cases (58% males, mean age (SD)=47.8(17.2)) as musicians, who we compared with 49,015 non-musician cases (60% males, mean age (SD)=47.5(17.4)), defined as absence of the above-listed musician keywords in their EHR, and matched for median age of record, sex, length of record, number of visits, race and ethnicity (Table 1).

257 medical diagnoses were found to be significantly associated with musician status (p-value<7.6 x 10^−6^, accounting for multiple testing) (Figure 2, Figure 3, for the full table see: Supplementary Table 3). Out of these, 242 diagnoses were associated with increased risk of diagnoses and 15 diagnoses were associated with decreased risk of diagnoses for the musician records. Among the top associations that were associated with increased risk of diagnoses were diseases of the larynx and vocal cords (OR=2.32, p<2.84 x 10^−191^), voice disturbance (OR=2.43, p<2.84 x 10^−191^), hearing loss (OR=1.36, p=5.98 x 10^−97^), pain in joint (OR=1.26, p=1.09 x 10^−125^), anxiety disorder (OR=1.25, p=7.67 x 10^−71^), sleep disorders (1.23, p=1.55 x 10^−47^), and back pain (OR=1.17, p=1.85 x 10^−47^). Chronic laryngitis (OR=3.02) and voice disturbance (OR=2.43) had the largest effect sizes. We summarized the top association within each medical diagnosis group in Table 2. Among the top associations that were associated with decreased risk were coronary atherosclerosis (OR=0.91, p=1.77 x 10^−10^), respiratory failure/insufficiency/arrest (OR=0.89, p=1.92 x 10^−09^), renal failure (OR=0.93, p=8.13 x 10^−07^), and ischemic heart disease (OR=0.92, p=1.65 x 10^−09^).

**Table 2.** Top specific diagnostic associations with musician status per phecode

| Group | Diagnosis | OR | p-value |
| --- | --- | --- | --- |
| Respiratory | Voice disturbance | 2.43 | $<2.84 \times 10^{-191}$ |
| Musculoskeletal | Pain in joint | 1.26 | $1.09 \times 10^{-125}$ |
| Symptoms | Cervicalgia | 1.45 | $2.25 \times 10^{-184}$ |
| Sense organs | Hearing loss | 1.36 | $5.98 \times 10^{-97}$ |
| Digestive | GERD | 1.21 | $9.13 \times 10^{-62}$ |
| Mental disorders | Anxiety disorder | 1.25 | $7.67 \times 10^{-71}$ |
| Neurological | Sleep disorders | 1.23 | $1.55 \times 10^{-47}$ |
| Endocrine/metabolic | Vitamin D deficiency | 1.15 | $1.48 \times 10^{-18}$ |
| Dermatologic | Disturbance of skin sensations | 1.24 | $4.51 \times 10^{-29}$ |
| Genitourinary | Menopausal and postmenopausal disorders | 1.21 | $3.77 \times 10^{-23}$ |
| Circulatory | Palpitations | 1.18 | $1.34 \times 10^{-27}$ |
| Neoplasms | Benign neoplasm of skin | 1.24 | $4.18 \times 10^{-27}$ |
| Infectious diseases | Viral warts and HPV | 1.19 | $7.81 \times 10^{-12}$ |
| Injuries and poisonings | Sprains and strains | 1.14 | $7.93 \times 10^{-15}$ |

**Table 3.**
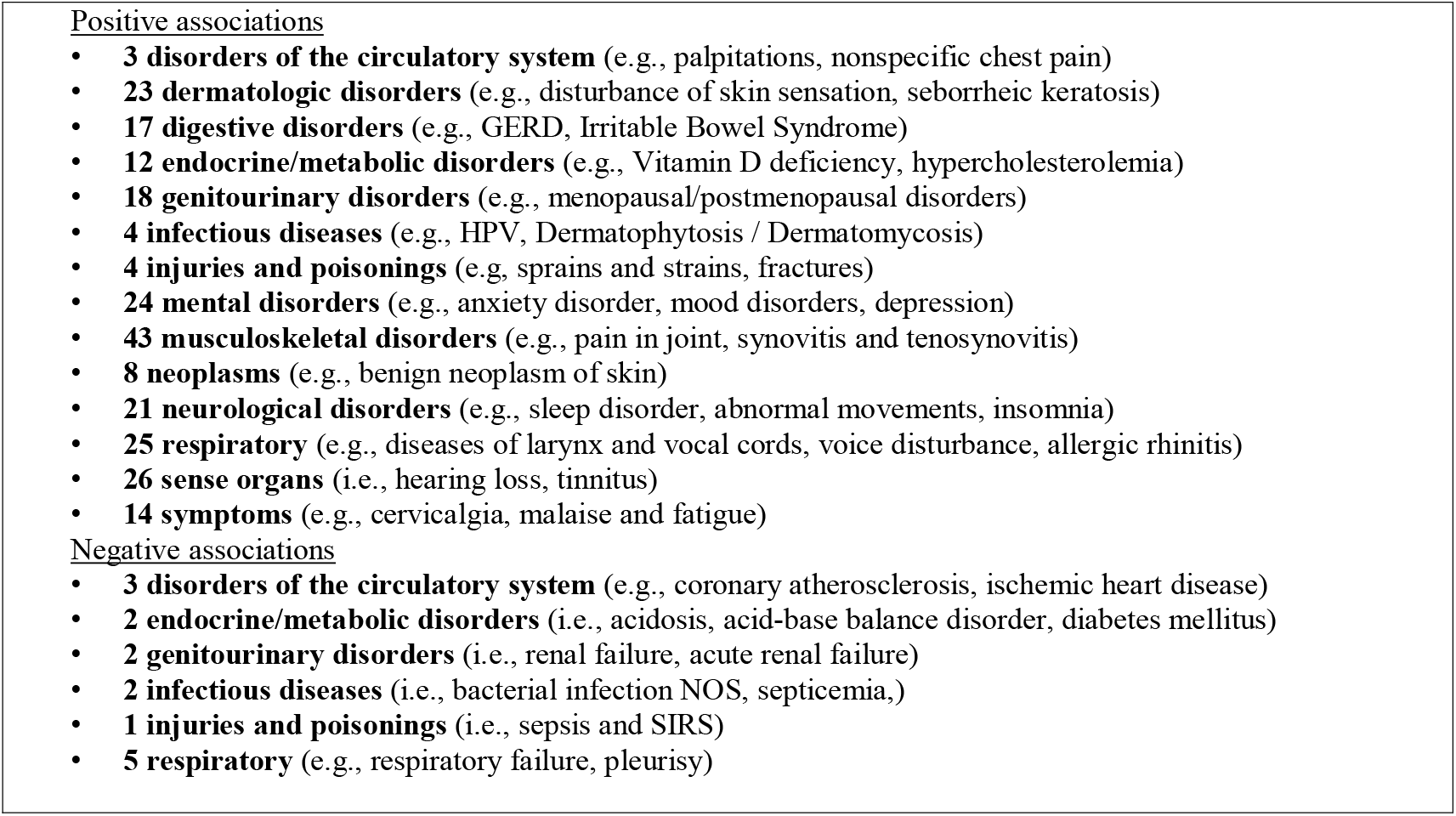
Summary of number of diseases within phecode categories

**Figure 2.**
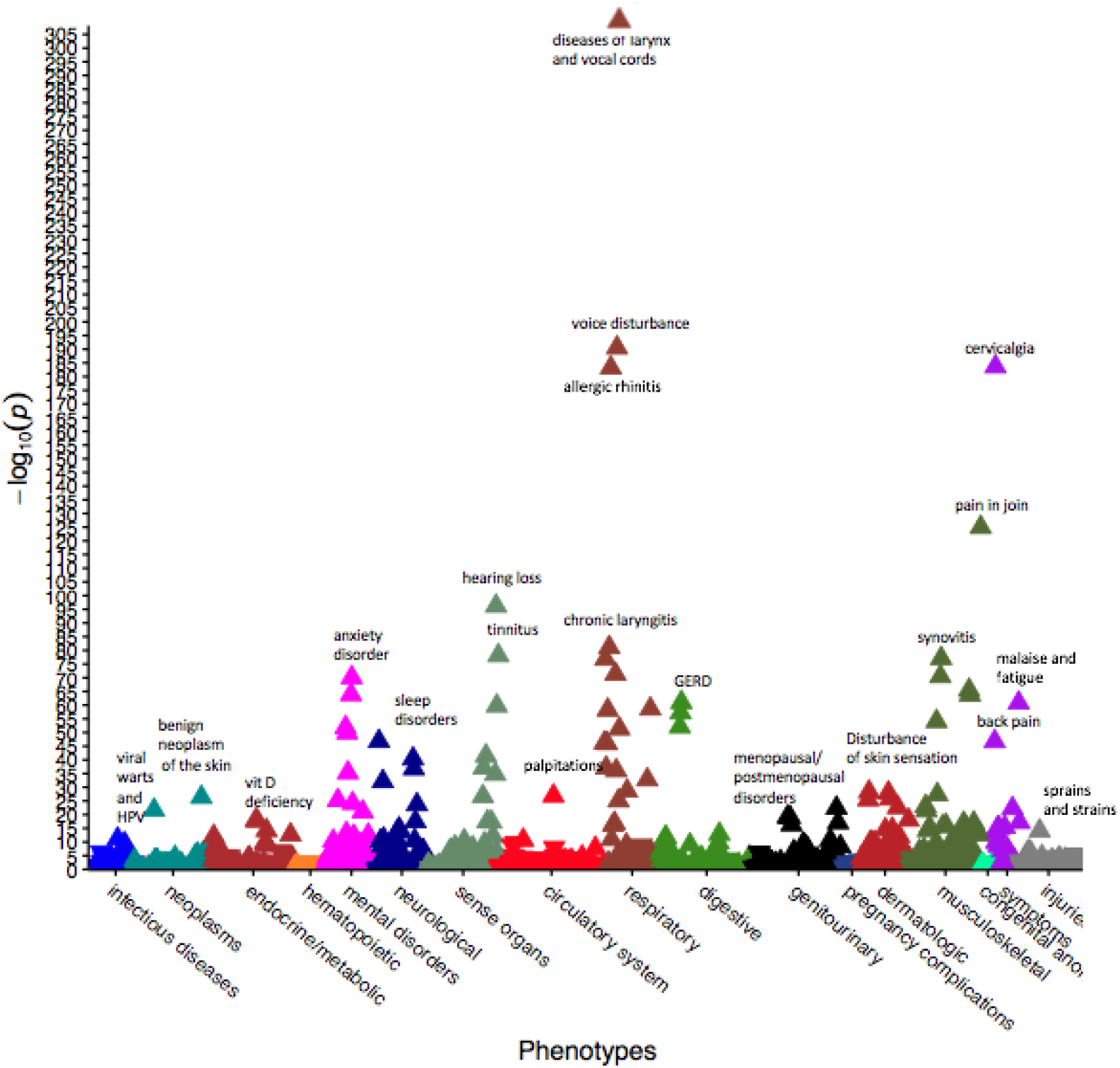
PheWAS plot of musicians versus control population in the Electronic Health Records. Phecode categories are on the x-axis, P value (-log_10_) on the y-axis, the triangles are the specific phecodes within the phecode categories. If the direction of the effect is positive, the arrows point upwards, if the direction of the effect is negative, the arrows point downwards.

**Figure 3.**
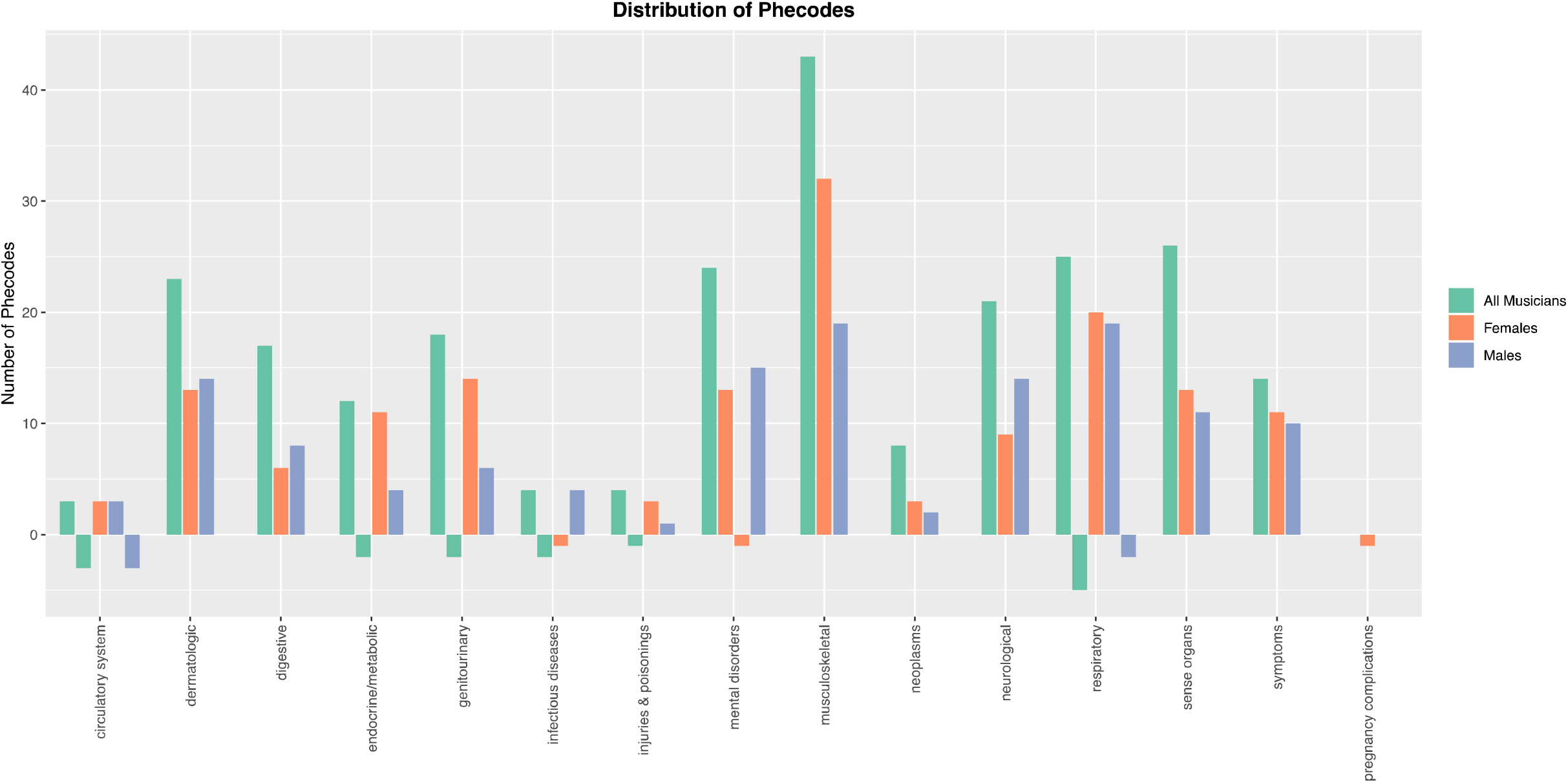
Phecode categories significantly associated with musicians

When we restricted the analyses to males, there were 135 diagnoses that were significantly associated with musician status (p-value<7.6 x 10^−6^) (Figure 3, Supplementary Table 4). Among those diagnoses, 130 associations were related to increased risk for diagnoses. The top associations related to increased risk for diagnoses were similar to the whole sample including voice disturbance (2.20, p=7.71 x 10^−237^), tinnitus (1.60, p=2.39 x 10^−55^), Generalized Anxiety disorder (OR=1.38, p=7.09 x 10^−24^), mood disorders (OR=1.23, p=1.83 x 10^−44^), and cervicalgia (OR=1.38, p=2.78 x 10^−68^). There were also 5 diagnoses that were associated with decreased risk including ischemic heart disease (OR=0.92, p=2.89 x 10^−08^), coronary atherosclerosis (OR=0.91, p=1.87 x 10^−08^), and congestive heart failure/non-hypertensive (OR=0.90, p=1.07 x 10^−06^).

Analyses in females identified 154 diagnoses that were significantly associated with musician status (p-values<7.6 x 10^−6^) (Figure 3, Supplementary Table 4). Among these, 151 diagnoses were related to increased risk of diagnoses including voice disturbance (OR=2.71, p=1.46 x 10^−304^), tinnitus (OR=1.53, p=2.72 x 10^−26^), throat pain (OR=1.52, p=1.01 x 10^−12^), and cervicalgia (OR=1.53, p=1.98 x 10^−117^). There were 3 diagnoses that showed decreased risk among female musicians including tobacco use disorder (OR=0.85, p=5.33 x 10^−08^), known or suspected fetal abnormality affecting management of mother (OR=0.87, p=7.56 x 10^−06^), and septicaemia (OR=0.83, p=1.55 x 10^−06^).

To examine whether the difference in the ORs between males and females was significantly different, we performed an interaction test (Sex * Musician/non-musician; Supplementary table 4). Voice disturbance, tinnitus, pain in joint as well as a number of other diagnoses were significantly more common among female musicians. On the other hand, mental health disorders, including anxiety disorder, depression, bipolar disorder, alcohol-related disorders and substance addiction and disorders were significantly more common among male musicians. Thus, after accounting for the different baseline prevalence of these conditions between males and females, there remains a significant difference in the strength of association between male and female musicians.

### String instrument players

We stratified to 3,209 players of string instruments (75% males) vs 16,045 (76% males) controls were (Table 1). 148 diagnoses were significantly associated with players of strings instruments (p<7.6 x 10^−6^) (Supplementary Table 5, Supplementary Figure 1, Supplementary Figure 2). Among the top associations positively associated with string instrument players were diseases of larynx and vocal cords (OR=1.77, p=8.21 x 10^−74^), voice disturbance (OR=1.88, p=2.39 x 10^−78^), tinnitus (OR=1.49, p=7.32 x 10^−21^), synovitis and tenosynovitis (OR=1.68, p=1.13 x 10^−56^), stiffness of joint (OR=1.63, p=5.65 x 10^−47^), and fasciitis (OR=1.49, p=1.15 x 10^−15^).

### Percussionists

886 players of percussion instruments (84% males) and 4430 controls (85% males) were identified and included in the analyses (Table 1). 19 diagnoses were significantly associated with players of percussion instruments (p<7.6 x 10^−6^) (Supplementary Table 6, Supplementary Figure 1, Supplementary Figure 3). Tinnitus (OR=1.63, p=4.66 x 10^−12^), Generalized anxiety disorder (OR=1.53, p=2.60 x 10^−08^), asthma (OR=1.32, p=6.28 x 10^−08^) and cervicalgia (OR=1.30, p=3.04 x 10^−08^) were among the top associations.

### Keyboard players

1,870 players of keyboards (35% males) and 9350 controls (34% males) were included in the analyses (Table 1). 61 diagnoses were associated with players of keyboard instruments (p<7.6 x 10^−6^) (Supplementary Table 7, Supplementary Figure 1, Supplementary Figure 4). Among these, 60 diagnoses were positively associated with keyboard players including voice disturbance (OR=1.64, p=6.21 x 10^−35^), synovitis and tenosynovitis (OR=1.57, p=1.85 x 10^−31^), stiffness of joint (OR=1.63, 1.62 x 10^−35^), and tinnitus (OR=1.35, p=2.88 x 10^−08^). Coronary atherosclerosis was negatively associated with keyboard players (OR=0.87, p=4.42 x 10^−06^).

### Brass instrument players

430 brass players (73% males) vs. 2150 controls (75 males) were identified (Table 1). There were no significant associations (p<7.6 x 10^−06^) identified.

### Woodwind instrument players

We included 308 (22% males) players of woodwinds vs. 1,540 (22% males) controls in the analyses (Table 1). There were no significant associations (p<7.6 x 10^−06^) identified.

### Vocalists

2,387 vocalists (43% males) and 11,935 controls (44% males) were included in the analyses (Table 1). 62 diagnoses were associated with vocalists (Supplementary Table 10, Supplementary Figure 1, Supplementary Figure 7). As expected, among the top associations were diseases of the larynx and vocal cords (OR=4.87, p<1.10 x 10^−208^), chronic laryngitis (OR=6.64, p=1.80 x 10^−139^), and voice disturbance (OR=5.13, p<1.10 x 10^−208^). Other strong associations were throat pain (OR=2.28, p=1.62 x 10^−33^), hearing loss (OR=1.28, p=1.85 x 10^−14^), GERD (OR=1.57, p=3.00 x 10^−99^), cervicalgia (OR=1.75, p=5.88 x 10^−122^), and sleep disorders (OR=1.15, p=2.92 x 10^−06^).

### Musicians between the ages of 5-17

There were 4,576 cases of musicians between the ages of 5 and 17 years (mean age(SD)=12.7(3.5) years; 48% males) and 22,880 controls (mean age(SD)=13.1(3.6) years; 47% males). 114 diagnoses were associated with musician status (Supplementary Table 11, Supplementary Figure 8). Among the strongest associations were anxiety disorder (OR=1.32, p=6.46 x 10^−50^), depression (OR=1.26, p=3.29 x 10^−33^), eating disorder (OR=1.48, p=4.95 x 10^−23^), and voice disturbance (OR=1.70, p=5.96 x 10^−26^). The largest effect was observed for voice disturbance (OR=1.70, p=5.96 x 10^−26^), followed by anorexia nervosa (OR=1.56, p=2.13 x 10^−12^).

## Discussion

In a sample of 14,379 musician cases and 71,895 matched controls identified in an Electronic Health Record database, we conducted the first and largest study to date to identify medical diagnoses associated with musician patients in an EHR context. We first created and validated a novel musician phenotype identification method for use in EHR, which showed a positive predictive value of 93%. We then replicated previous associations of musician status with medical problems, including musculoskeletal problems^5^, skin disorders^6^, respiratory and hearing problems^7 2^, as well as mental health problems^17^, and also detected novel associations. Furthermore, we identified protective effects for a number of diagnoses, including coronary atherosclerosis and renal failure. These findings are in line with a study indicating lower blood pressure and heart rate in musicians, compared to controls^18^, as well as studies suggesting that active music engagement has similar training effects to physical exercise^18, 19^.

When we stratified our analyses by instrument family, results of health problems aligned with type of instrument. There was substantial overlap in diagnoses associated with different instrument families as well as distinct patterns of effects analogous to the physical demands of each instrument. For example, for vocalists, the strongest associations were diseases of the larynx, and vocal cords, voice disturbance, and chronic laryngitis. Players of percussion instruments demonstrated higher rates of hearing loss, while players of string instruments were more commonly treated for pain in joint and cervicalgia as well as voice disturbance. Keyboard players experienced higher rates of pain and stiffness of joint. These findings provide further face validity for the method of musician identification in the EHR and increase confidence in the novel associations presented.

When it comes to protective effects related to the family instruments, these were restricted to keyboard players and, as in the total sample of musicians, were related to less risk for cardiovascular diseases. Although statistical power could be a factor why we did not see such effects for brass, woodwind, and percussion instrument players, this does not seem to be the case for players of strings instruments, and vocalists, as these samples of individuals were larger than the keyboard players. It is possible that playing keyboards has greater cardiovascular benefit than playing string instruments or singing. The effect may be confounded by the music genre, years of training, or performance settings. Further studies are needed to better understand the underlying physiology at play and to replicate our findings.

We also found strong associations of musician status with a number of mental health disorders, some of which were already present in childhood and adolescence, including anxiety disorders, depression, post-traumatic stress disorder, and bipolar disorder, suggesting that musicians may benefit from mental health support integrated into musical training. The exact nature of the association between musical activities and mental health problems is unclear. A recent twin modelling study found that this association is more likely explained by shared genetic or environmental factors that affect both risk for mental health problems and musical achievement ^20^. Evidence for shared genetic or environmental susceptibility also comes from a study that found that higher polygenic scores for schizophrenia and bipolar disorder were associated with artistic society membership or creative profession^21^.

A previous study on 377 musicians reported that the chief complaints of musicians were mental health-related - performance-related anxiety, social anxiety, and depression - and were more common in female musicians^3^. We found that although the effect sizes of depression and anxiety-related disorders are slightly higher in males, such problems were strongly associated with both male and female musicians.

Our study included a total of 86,274 individuals. This unprecedented sample size provided us with the statistical power to be able to detect the prevalence of 257 diagnoses that were more common among musicians than non-musicians. Several of those diagnostic associations have not been previously described in the literature, including circulatory disorders, genitourinary disorders, and endocrine/metabolic disorders. Though speculative, a subset of these disorders may be associated with environmental risk factors associated with being a musician.

Apart from mental health disorders, our study identified a wide range of other disorders that healthcare professionals should be aware of, in order to provide treatment catered to the needs of musicians. Furthermore, our study demonstrates how information on a persons’ occupational and vocational history can provide useful insights in terms of preventing and anticipating the medical problems that may be related to a specific occupation^22^. Therefore, it is possible that the effect sizes for certain disorders that are directly related to a person’s ability to perform their music (e.g., voice disturbance), are higher than the true estimate, given that individuals may not seek treatment for issues that are less likely to affect their professional life. Future studies in the healthcare context could use EHRs to identify risks associated with other types of occupation. Finally, we found protective effects of being a musician, including lower risk for coronary atherosclerosis and renal failure, providing further evidence for potential cardiovascular benefits associated with the increased physiological demands associated with playing a musical instrument.

Despite the enormous power that research in the EHR provides, there are certain limitations of the current approach. We were not able to separate from the records whether an individual is an amateur vs. a professional musician, how many years of training they had, whether they are part of an ensemble or solo musician, or the music genre. Although we sometimes had information on the type of instrument, this was not always the case, as we would often encounter charts reporting ‘musician’ as the only relevant keyword. Also, future studies can examine whether there are differences in health problems between single-instrumentalists and multi-instrumentalists. There is also the possibility that a physician is more likely to identify a patient as a musician if their chief complaint during a clinic encounter is related to their music. The impact of this in our results could be differential loss of power across diagnoses, that would in turn make it difficult to compare the effect sizes between different phenotypes. However, it is noteworthy that in our EHR, each individual does not represent only one phenotype, but many phenotypes that they accumulated throughout their medical record. This allows us to investigate a range of phenotypes that are not necessarily attributed to musician status. Finally, VUMC has specialty clinics, including a voice clinic, that attracts individuals from across the country, and therefore, such specialty clinics may enrich for different diagnoses.

## Conclusions

We conducted the first PheWAS of musicians using an EHR framework in over 80,000 individuals and found both risk and protective associations with many medical conditions. These included medical complaints involving the musculoskeletal system, the respiratory, endocrine, and metabolic systems, as well as mental health problems. On the other hand, we also identified a number of protective effects by observing diagnoses such as cardiovascular disease, respiratory failure, and renal failure, that were less common in musicians than in controls, in line with literature indicating that active musical engagement has similar health benefits to athletic engagement. Our study highlights the need for management, prevention, and health education of musicians and the health care providers treating them. Finally, this study serves as a proof of principle to demonstrate that EHR studies, and PheWAS in particular, can be effectively used to identify medical conditions associated with specific occupations.

## Data Availability

Due to data sharing restrictions related to privacy concerns in the EHR, the datasets generated from our hospital population will not be publicly available, however, all scripts used in the study are available upon request.

## Acknowledgements

This project was supported by funding from the National Institutes of Health, NIH Common Fund through the Office of the NIH Director under Award Number DP2HD098859. The content is solely the responsibility of the authors and does not necessarily represent the official views of the National Institutes of Health. We would also like to thank the Nashville Musicians Association and especially Kathy Osborne for providing us with the list of the most popular musical instruments.

## Synthetic Derivative

The project described was supported by the National Center for Research Resources, Grant UL1 RR024975-01, and is now at the National Center for Advancing Translational Sciences, Grant 2 UL1 TR000445-06. The content is solely the responsibility of the authors and does not necessarily represent the official views of the NIH.

