## Supplementary figures for "The medical signature of musicians: A Phenome-wide association study using an Electronic Health Record database"

### Slide 1
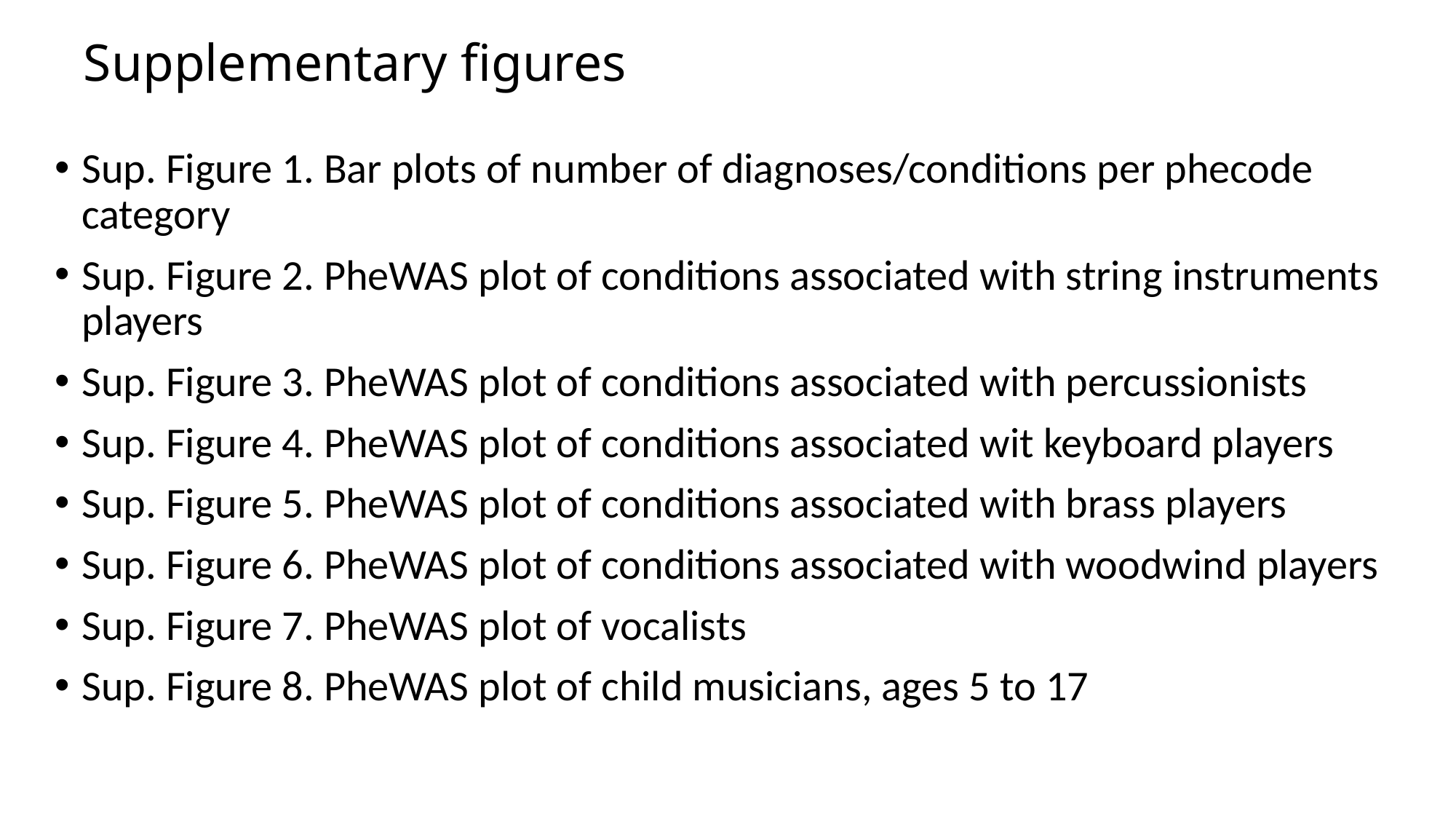

#### Supplementary figures
Sup. Figure 1. Bar plots of number of diagnoses/conditions per phecode category
Sup. Figure 2. PheWAS plot of conditions associated with string instruments players
Sup. Figure 3. PheWAS plot of conditions associated with percussionists
Sup. Figure 4. PheWAS plot of conditions associated wit keyboard players
Sup. Figure 5. PheWAS plot of conditions associated with brass players
Sup. Figure 6. PheWAS plot of conditions associated with woodwind players
Sup. Figure 7. PheWAS plot of vocalists
Sup. Figure 8. PheWAS plot of child musicians, ages 5 to 17

### Slide 2
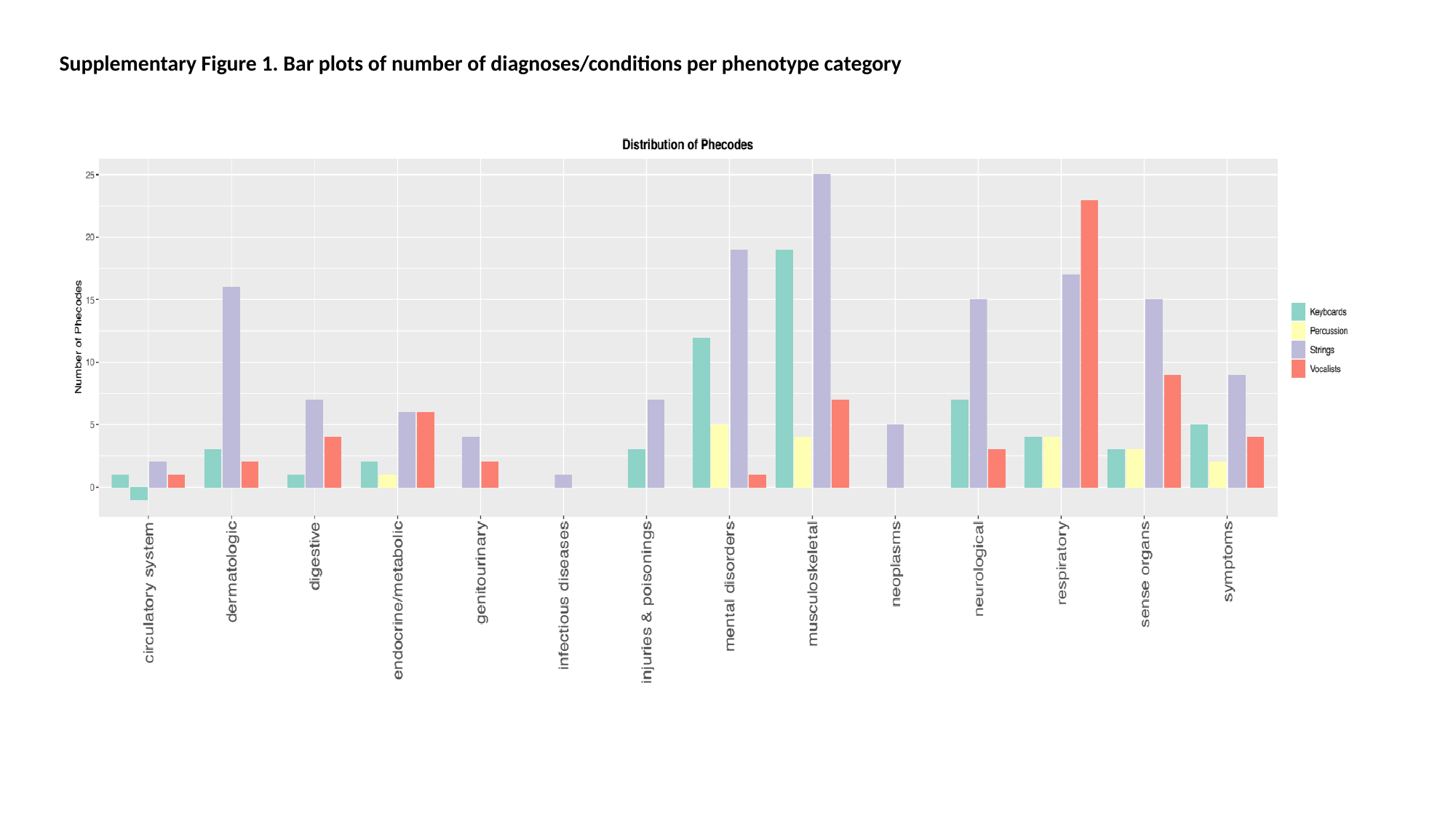

#### Supplementary Figure 1. Bar plots of number of diagnoses/conditions per phenotype category

### Slide 3
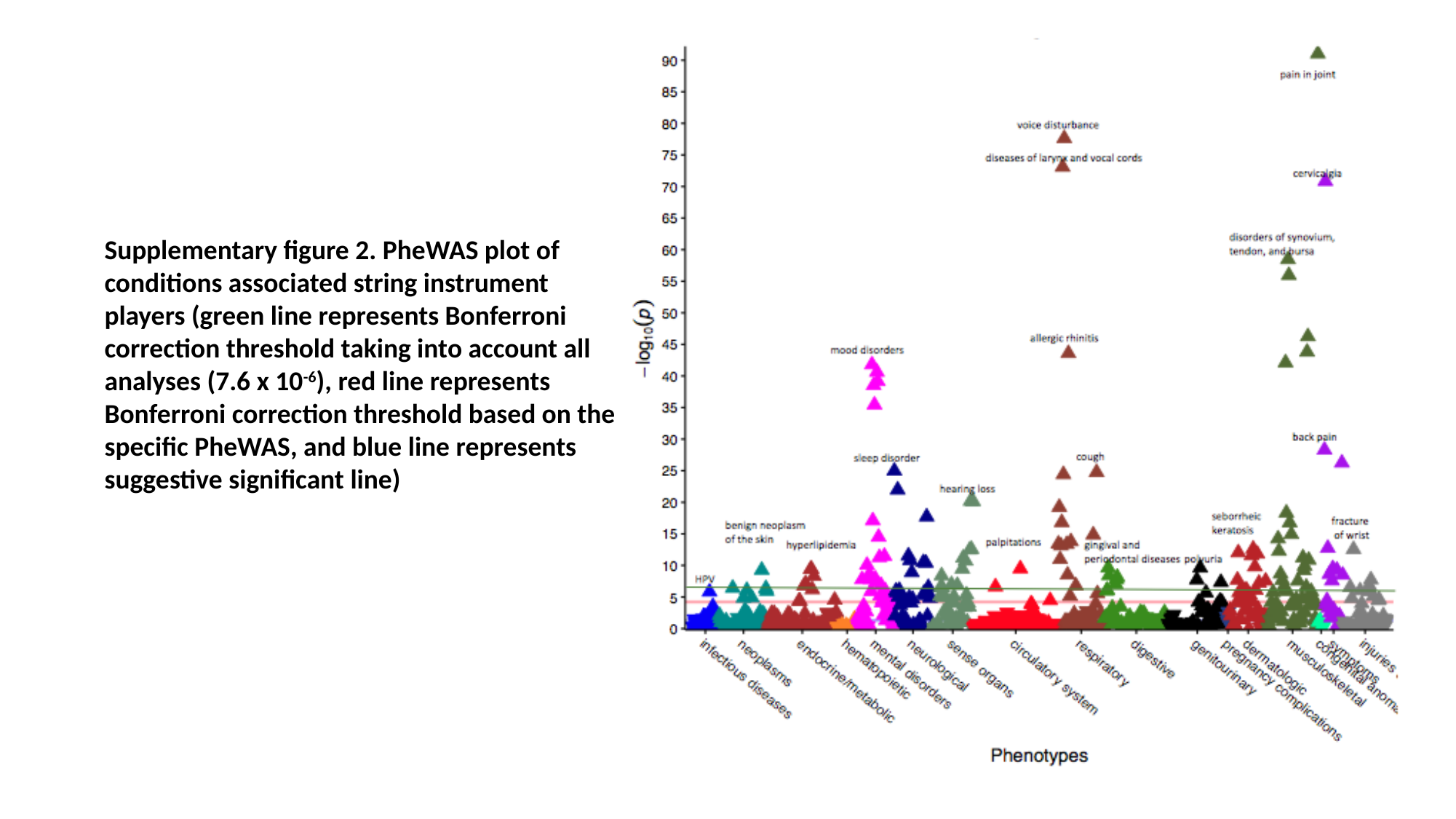

Supplementary figure 2. PheWAS plot of conditions associated string instrument players (green line represents Bonferroni correction threshold taking into account all analyses (7.6 x 10^-6^), red line represents Bonferroni correction threshold based on the specific PheWAS, and blue line represents suggestive significant line)

### Slide 4
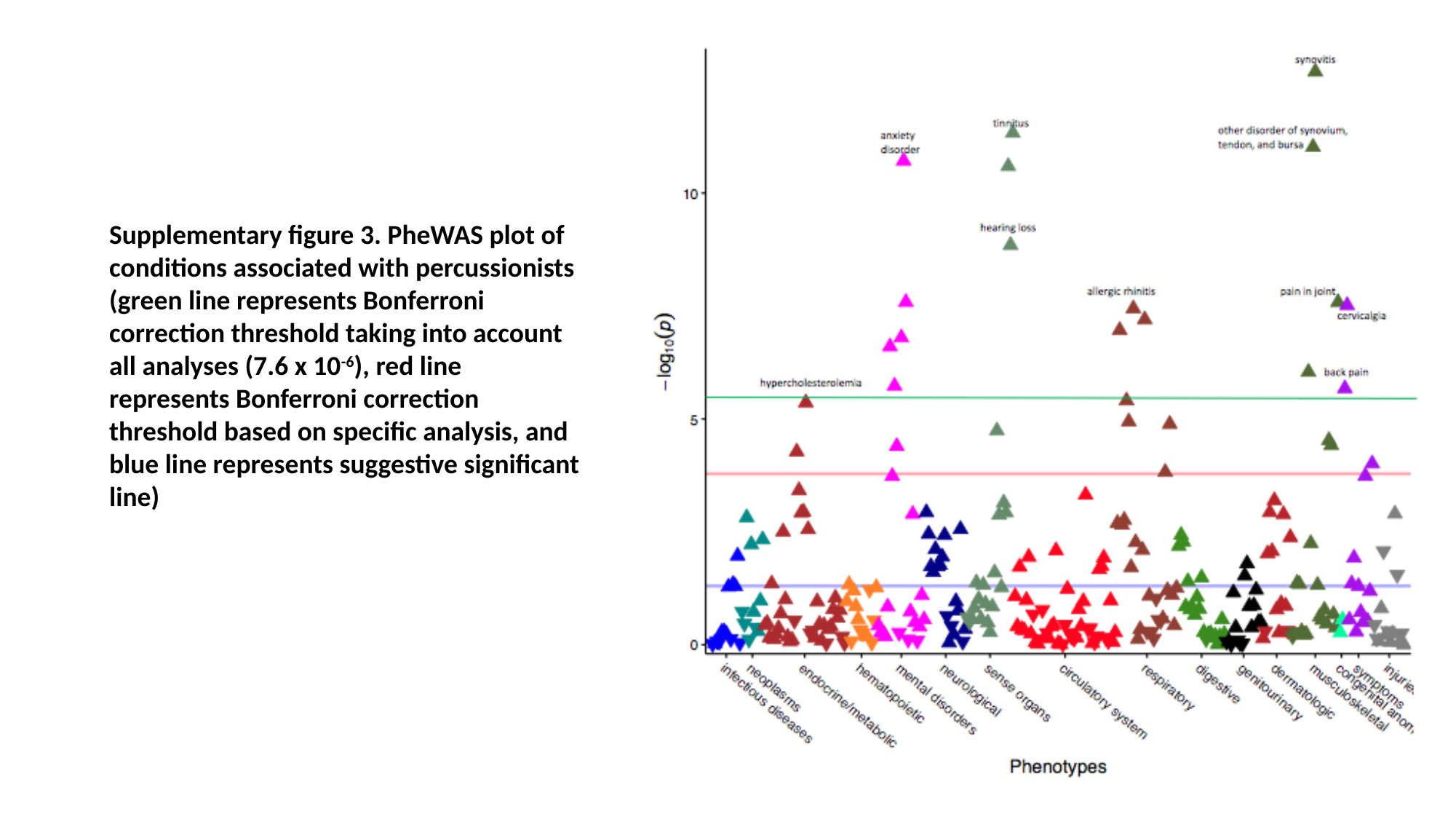

Supplementary figure 3. PheWAS plot of conditions associated with percussionists (green line represents Bonferroni correction threshold taking into account all analyses (7.6 x 10^-6^), red line represents Bonferroni correction threshold based on specific analysis, and blue line represents suggestive significant line)

### Slide 5
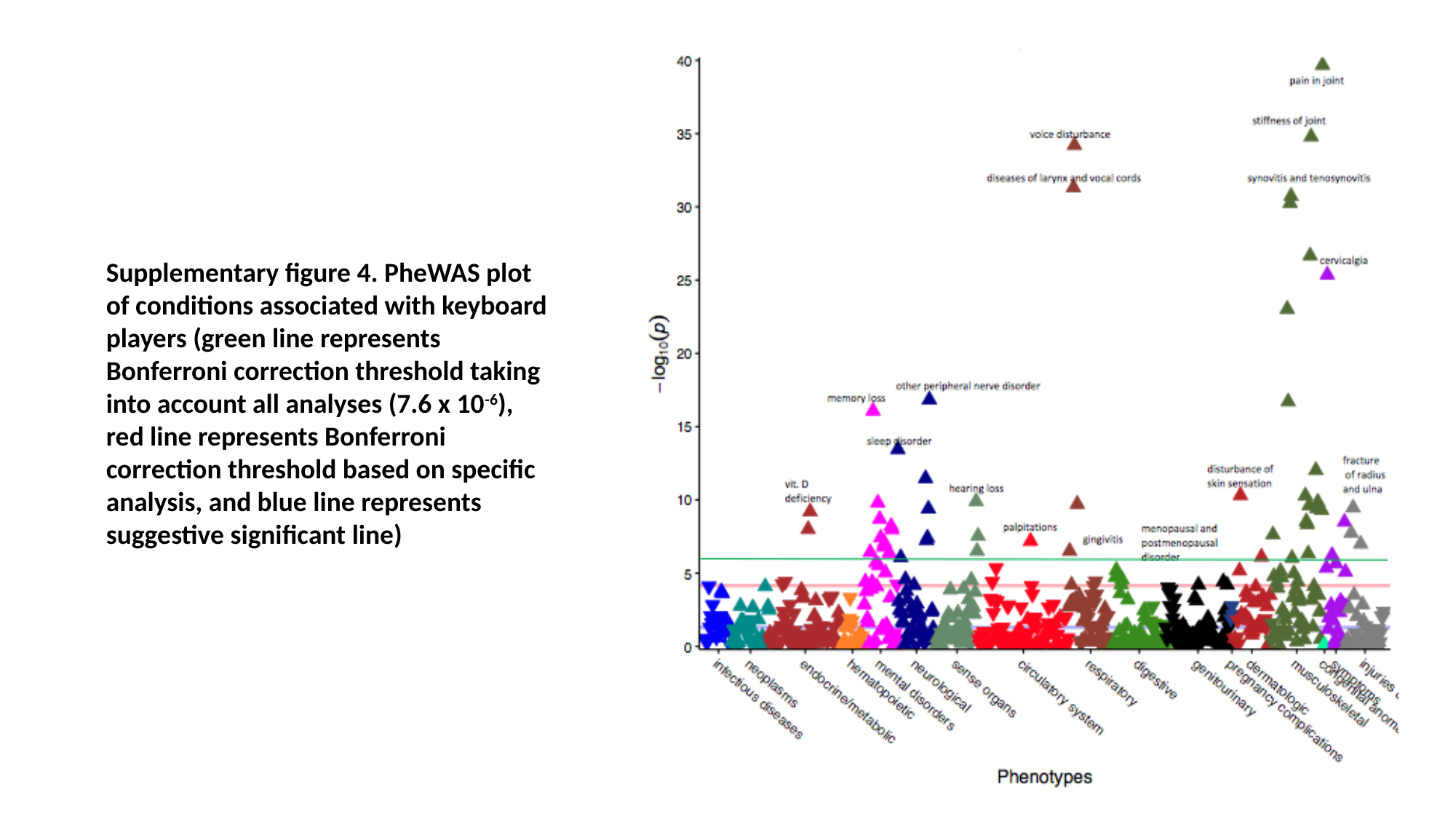

Supplementary figure 4. PheWAS plot of conditions associated with keyboard players (green line represents Bonferroni correction threshold taking into account all analyses (7.6 x 10^-6^), red line represents Bonferroni correction threshold based on specific analysis, and blue line represents suggestive significant line)

### Slide 6
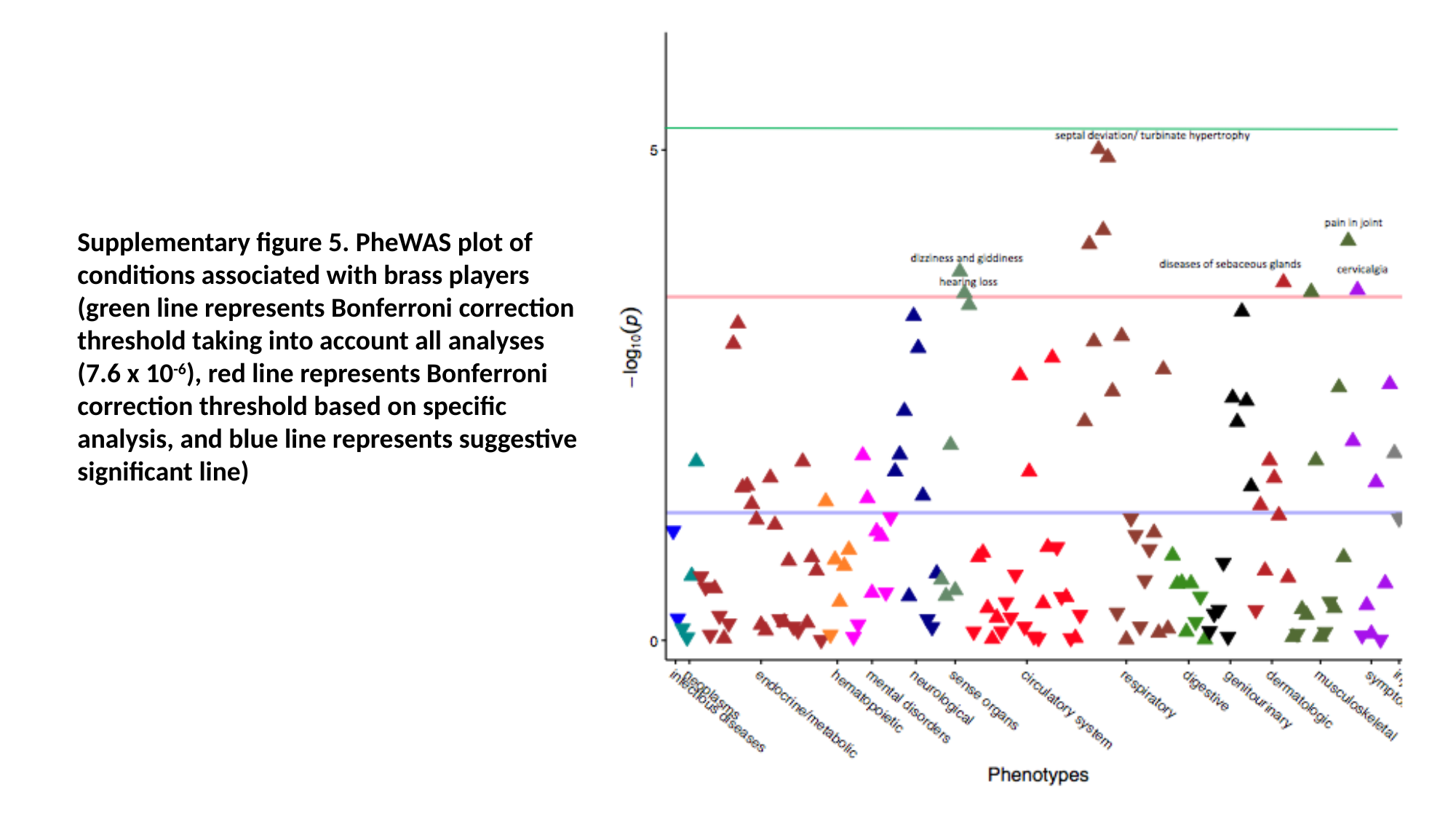

Supplementary figure 5. PheWAS plot of conditions associated with brass players (green line represents Bonferroni correction threshold taking into account all analyses (7.6 x 10^-6^), red line represents Bonferroni correction threshold based on specific analysis, and blue line represents suggestive significant line)

### Slide 7
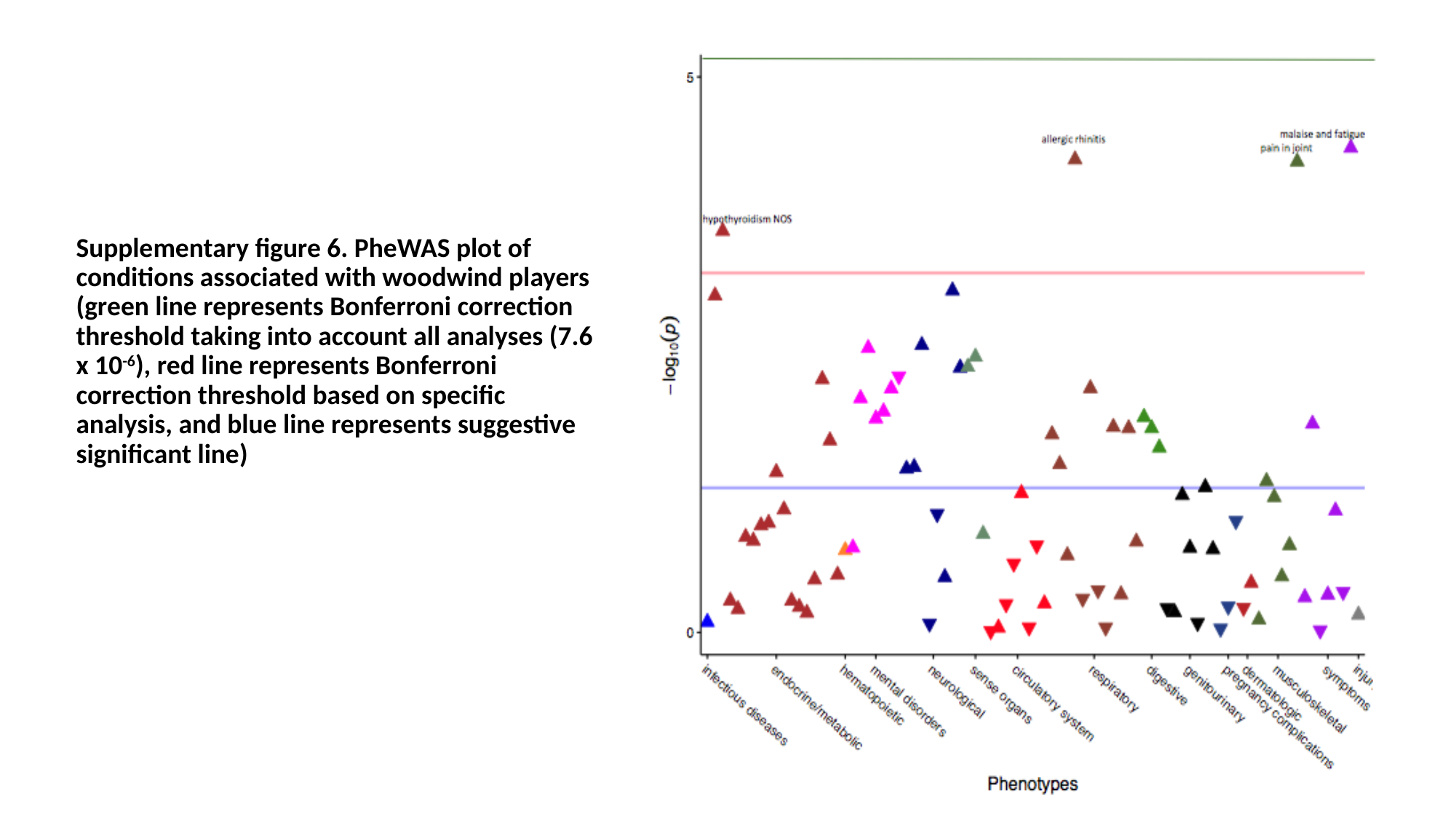

#### Supplementary figure 6. PheWAS plot of conditions associated with woodwind players (green line represents Bonferroni correction threshold taking into account all analyses (7.6 x 10^-6^), red line represents Bonferroni correction threshold based on specific analysis, and blue line represents suggestive significant line)

### Slide 8
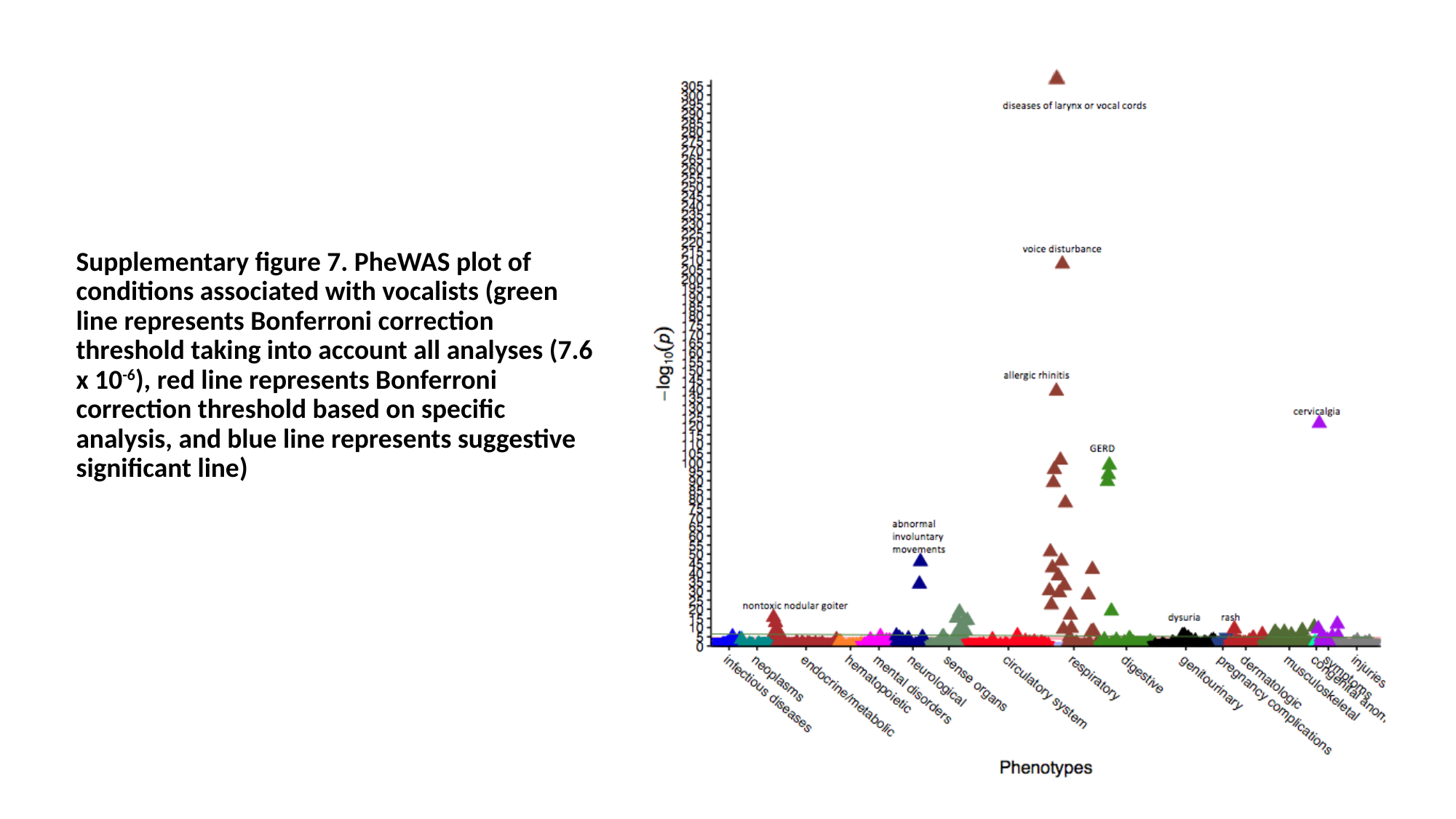

#### Supplementary figure 7. PheWAS plot of conditions associated with vocalists (green line represents Bonferroni correction threshold taking into account all analyses (7.6 x 10^-6^), red line represents Bonferroni correction threshold based on specific analysis, and blue line represents suggestive significant line)

### Slide 9
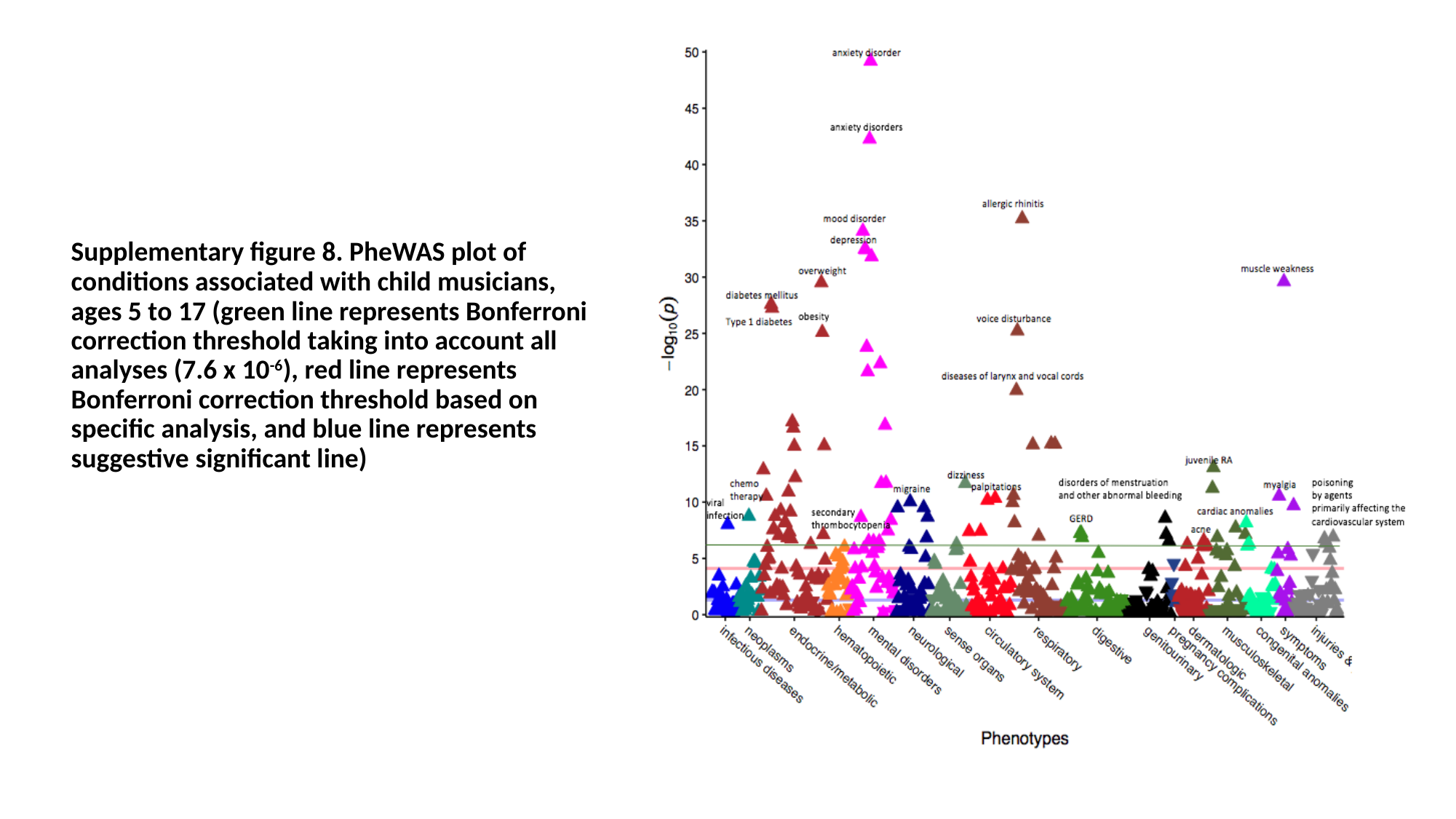

#### Supplementary figure 8. PheWAS plot of conditions associated with child musicians, ages 5 to 17 (green line represents Bonferroni correction threshold taking into account all analyses (7.6 x 10^-6^), red line represents Bonferroni correction threshold based on specific analysis, and blue line represents suggestive significant line)
